# COVID-19 in hospitalized patients in 4 hospitals in San Isidro, Buenos Aires, Argentina

**DOI:** 10.1101/2021.07.30.21261220

**Authors:** María Ana Barra, Nelson Adolfo Carlos Medinacelli, Camilo Andres Meza Padilla, Lucrecia Di Rocco, Ramiro Manuel Larrea, Giuliano Gaudenzi, Verónica Vanesa Mastrovincenzo, Elisa Raña, Inés Moreno, Dagmar Ana Sörvik, Andrea Sarlingo, Florencia Dadomo, Miguel Torrilla

## Abstract

In December 2019, a novel illness called coronavirus disease 2019 (COVID-19) was described in China and became pandemic in a few months. The first case was detected in Argentina on March 3, 2020.

A multicentre prospective observational cohort study on hospitalized patients with COVID- 19 was conducted in 4 hospitals in San Isidro district from March 1, 2020 to October 31. Data was obtained by the attendant physician. 668 patients were included, the median age was 54 years, and 42.7% were female. Male sex and older age were associated with COVID-19 disease and more strongly with severity. Most frequent symptoms were fever and cough followed by dyspnoea, myalgia, odynophagia, headache, anosmia, and diarrhoea. Non-severe patients had more upper respiratory symptoms while severe patients had mainly lower respiratory symptoms on admission. Most common comorbidities were arterial hypertension, diabetes, and cardiovascular disease. A great proportion of patients had normal thorax X-ray and ground-glass opacity in tomography. In severe patients, radiography and tomography had a predominant ground – glass pattern, but normal radiography and tomography on presentation were present in 22% and 5.9%, respectively. The absence of fever and normal radiology on admission neither excluded the disease nor further severity. PCR elevation was related with COVID-19 disease and with severity, while lymphopenia was more related with the disease and leukocytosis and thrombocytopenia with severity. 8, 4% of patients were health care workers. The mortality rate was 12.4%, 32.7% in severe patients and 61.2% in ventilated patients. Mortality was higher in the public hospital, probably associated with patients with older age and more comorbidities. All these observations can contribute to the knowledge of this disease in terms of diagnosis and prognosis.

## Introduction

In December 2019, a novel coronavirus was identified, called severe acute respiratory syndrome coronavirus 2 (SARS-CoV-2), causing a disease named coronavirus disease 2019 (COVID-19). This novel disease became a pandemic in a short time and caused thousands of deaths in many countries. Its clinical characteristics, risk factors, and severity have been described in some recent publications around the world. The first case in Argentina was detected on March 3, 2020. This study aims to describe the clinical characteristics, risk factors, severity, and outcome of hospitalized patients with COVID-19 in 4 hospitals in San Isidro district, Buenos Aires, Argentina, and to contribute with our findings to the knowledge of this disease.

## Methods

This is an observational, prospective, multicentre cohort study conducted in 4 hospitals in San Isidro, Buenos Aires, Argentina. The first cases were obtained retrospectively to include all patients hospitalized with COVID-19 from March 1, 2020, and were included patients admitted until August 31,2020 and followed until discharged or deceased. The recruitment ended days earlier in some of the hospitals due to the overwhelming task of the attending physicians. Patients were confirmed by a reverse transcriptase–polymerase-chain reaction (PCR-RT) for SARS-COV-2 assay of a specimen obtained with a nasopharyngeal swab. Patients included were older than 14 years old admitted to any of the 4 hospitals. Participant hospitals were Hospital Central San Isidro, a public hospital, and 3 private hospitals: Sanatorio Trinidad San Isidro, Sanatorio San Lucas and Sanatorio Las Lomas. Most patients admitted in the public hospital have lower incomes and no health security. To ensure isolation, many patients with mild COVID were transferred to isolation centers, while in private hospitals patients with mild COVID were admitted.

All data have been provided by attending physicians and registered from the day respiratory samples were obtained until patients were discharged. Data included clinical symptoms, epidemiological characteristics, comorbidities, laboratory, radiology, treatment, and clinical outcome. Initial laboratory testing was defined as the first test results available, typically within 24 hours of admission. For initial laboratory testing and clinical studies for which not all patients had values, the percentages of total patients with completed tests are shown. Fever was considered axillary temperature equal or more than 37,5°C. Comorbidities considered were: diabetes, arterial hypertension, use of ACE inhibitors or ARBs, chronic renal failure, cardiovascular disease, asthma, COPD, structural lung disease, oncohemathological disease, smoking, chronic liver disease and immunosuppressive drugs. Obesity was registered as an observation as it was estimated and could not be measured by the attendant physician.

Categorical variables were processed by number of cases and proportions, and included sex, symptoms, radiology, comorbidities, condition at hospital, discharge, severity, and treatment. Severity was considered when any of the following was present: transfer to a closed unit, oxygen saturation lower than 93, increase of more than 50% in lung infiltrates in less than 48 hours, impaired consciousness, CURB greater than 1, assisted ventilation, hemodynamic instability. For continuous variables, median and IQR was calculated and included age, temperature before admission, and laboratory parameters on admission.

Clinical outcomes were monitored until October 31, the final date of follow-up. No sample size calculation was performed; the sample size was established by the time window of the study. All analyses were performed with the use of R software (R Project for Statistical Computing; R Foundation). No analysis for statistical significance was performed given the descriptive nature of the study.

The study was approved by the Institutional Evaluation Committee of the Austral Hospital registered in the Office of Human Research Protections IORG0006075, IRB00007319 – Universidad Austral IRB and in the Central Buenos Aires Province Ethic Committee. Informed consent was obtained verbally owing to the contagiousness of the disease and for cases in which it could not be obtained it was waived as it is a low risk study and all data were analyzed anonymously. No patient denied inclusion in the study.

## RESULTS

### DEMOGRAPHIC AND CLINICAL CHARACTERISTICS

668 cases of COVID-19 were registered in the 4 institutions in the established period of time (Supplementary material Table 1-Cases classified by hospitals). The first case was confirmed on March 12, 2020 at Sanatorio San Lucas with first symptoms on February 29. The median age was 54 years, most frequently between 50 and 60 years (19.6%) with a predominance of men (57.3%) (Table1). The median age of the non-severe cases was 52 years with 20.3% between 40 and 50 years, of the severe was 61 years, and of the deceased patients was 77 years (70-87) years. The male percentage was 52.9% in non-severe cases, 70.2% in the severe, and 66.3% in the deceased. The most frequent symptoms were fever (66.9%) and cough (57%), followed by dyspnoea (30.7%), myalgia (25.3%), odynophagia (21%) and headache (18.6%). Less frequent were anosmia (12.7%), diarrhoea (7.5%), and fatigue (6.9%). Dyspnoea was present in 21.7% of non-severe patients, 56.7% of the severe, and in 41 deceased patients (49.4%). Odynophagia was present in 21.7% of non-severe patients and in 18.7% of severe cases and 10.8% of deceased. Only two of the dead patients had anosmia (2.4%), while 13.9% of the non-severe had.

**TABLE 1 –.**
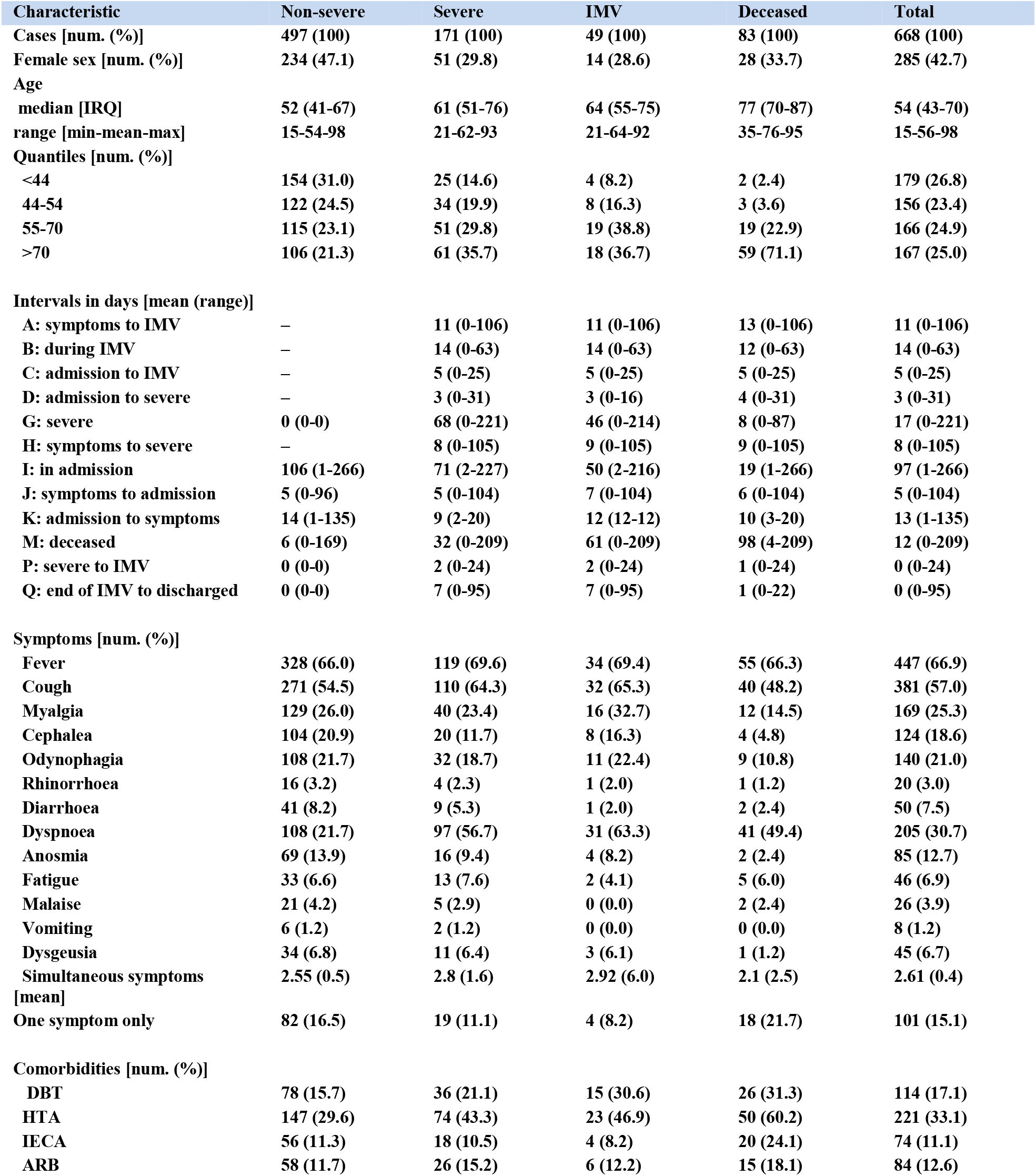
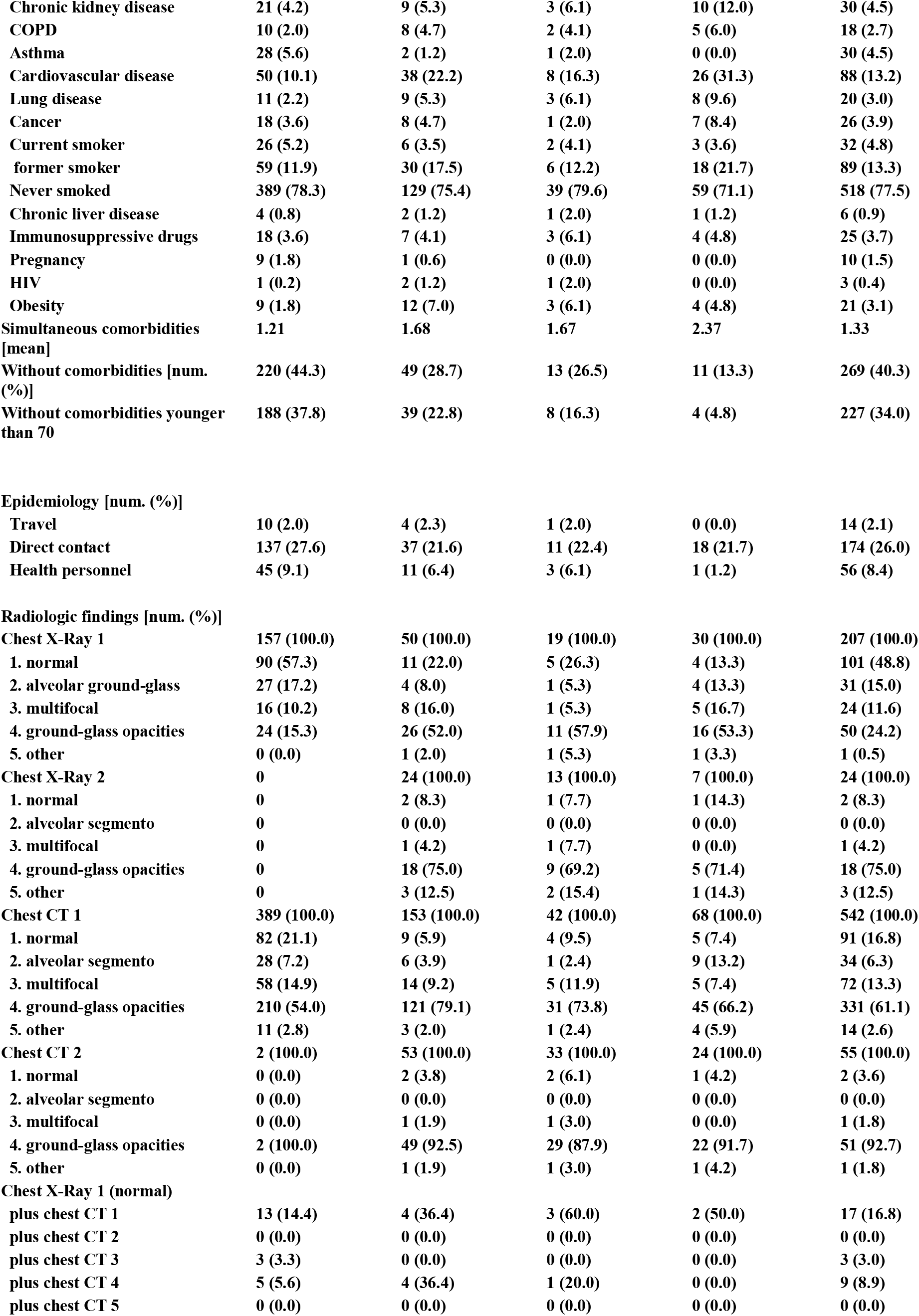
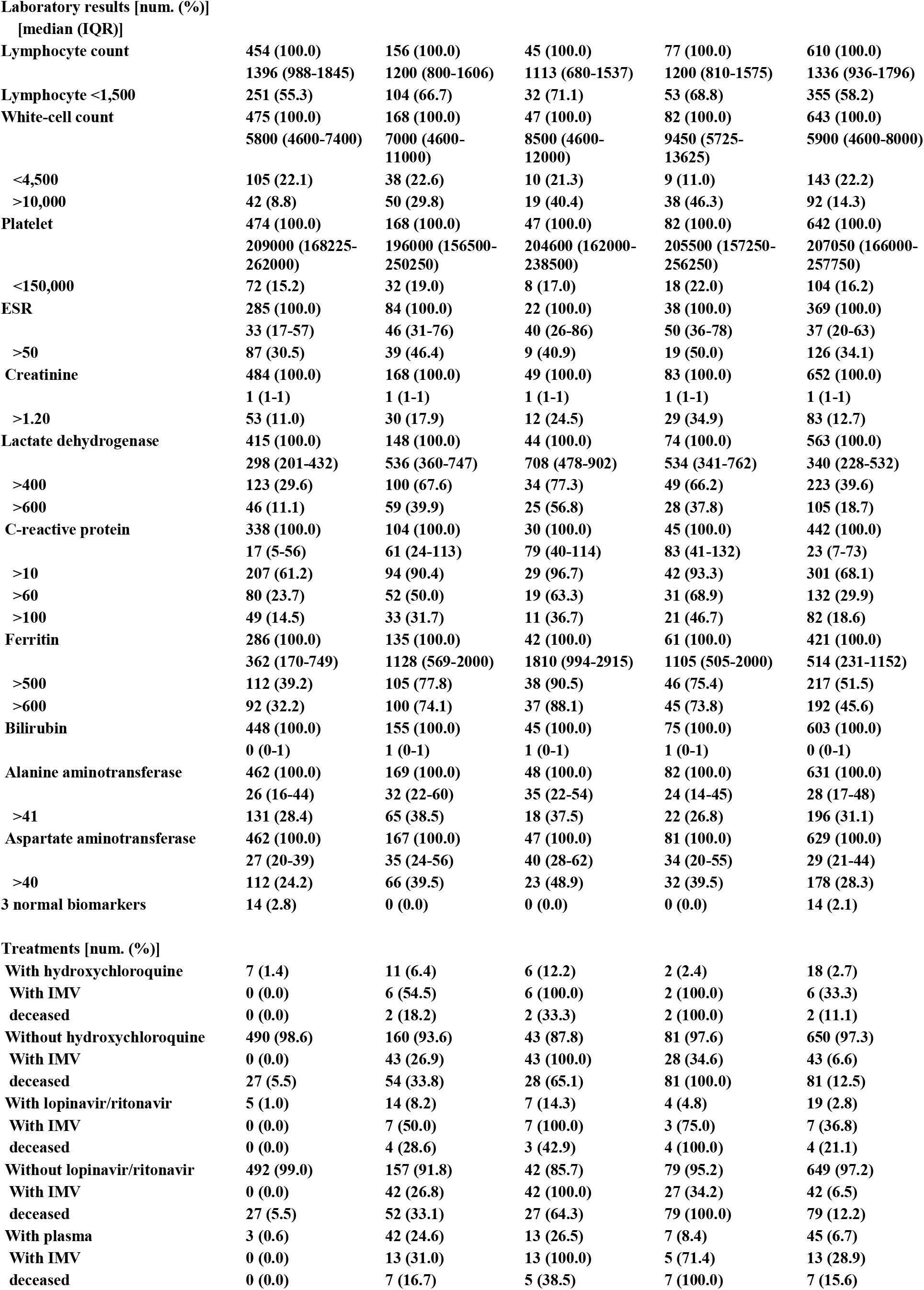
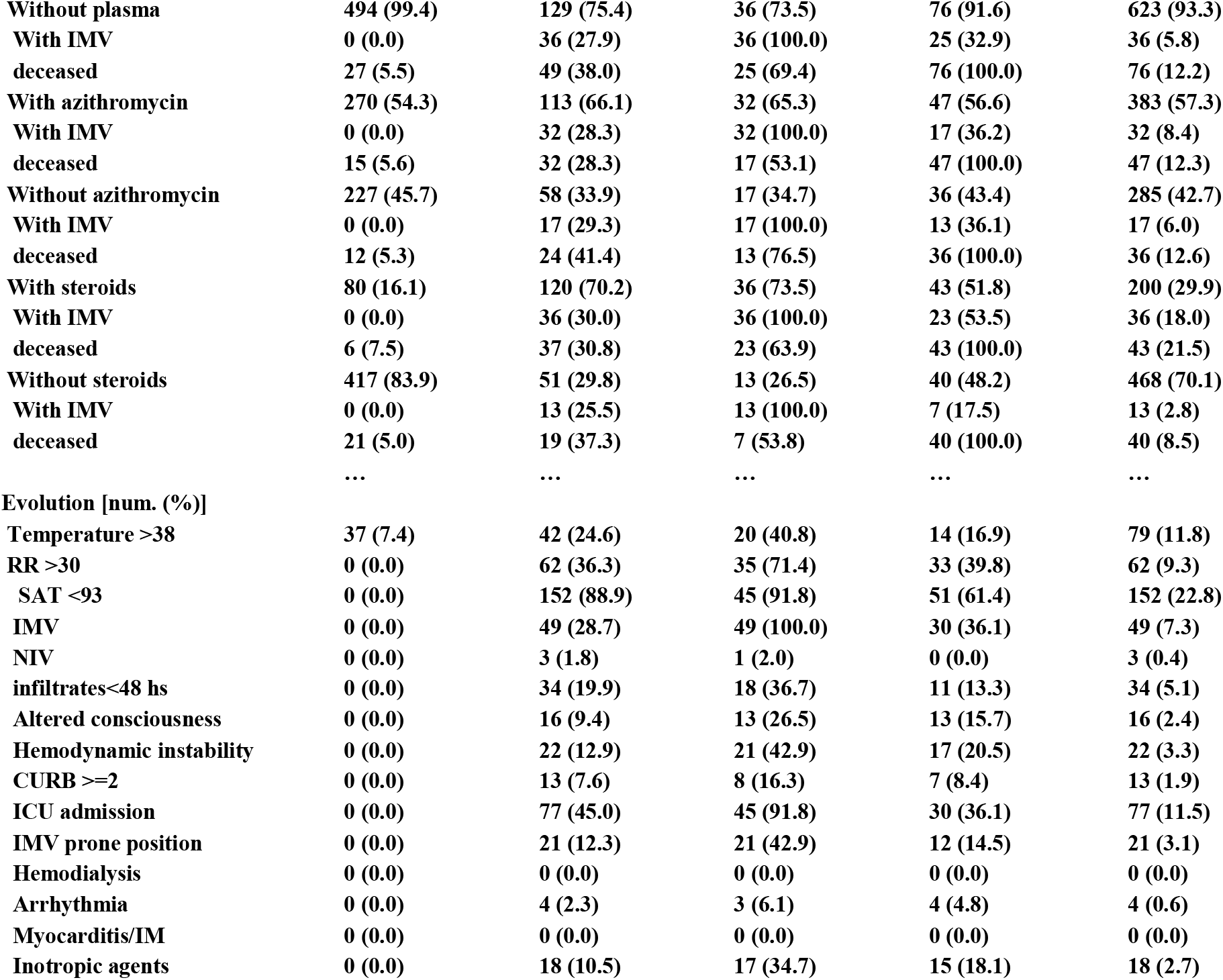
Cases classified by outcome.

The mean of simultaneous symptoms was 2.61 with 15.1% of patients with only one symptom. Severe patients had 2.8 simultaneous symptoms, 2.55 in non severe and 2.1 in deceased.

### COMORBIDITIES

Most frequent comorbidity was hypertension in 221 patients (33.1%), followed by diabetes in 114 patients (17.1%) and cardiovascular disease in 88 patients (13.2%). Most patients (77.5%) have never smoked. Less frequent were asthma (4.5%), chronic kidney disease (4.5%), obesity (3.1%) (probably underestimated), cancer (3.9%), COPD (2.7%) and use of immunosuppressive drugs (3.7%). 29.6% of the non-severe cases were hypertensive, 43.3% of severe and 60.2% of the dead. Other comorbidities were also more frequent in severe disease: COPD (4.7 vs. 2.0% in non-severe), cardiovascular disease (22.2 vs. 10.1%), immunosuppressive drugs (4.1 vs. 3.6%) and diabetes (21.1 vs. 15.7%) while asthma was more frequent in non severe (5.6 vs. 1.2% in severe). In patients who died, hypertension, diabetes, and cardiovascular disease were the most frequent comorbidities.

40.3% of patients did not have comorbidities. The mean of simultaneous comorbidities was 1.33.The mean of simultaneous comorbidities was 1.21 in no severe patients, 1.68 in severe patients, and 2.37 in dead patients. 44.3% of non-severe patients had no comorbidities, 28.7% of the severe, and 13.3% of dead. 4.8% of deceased patients had no comorbidities and were younger than 70 years old.

36.5% of the patients had increased epidemiological risk with 174(26%) patients who had known contact with confirmed COVID-19 patients and 56 patients (8.4%) were health personnel.

**Figure 1.**
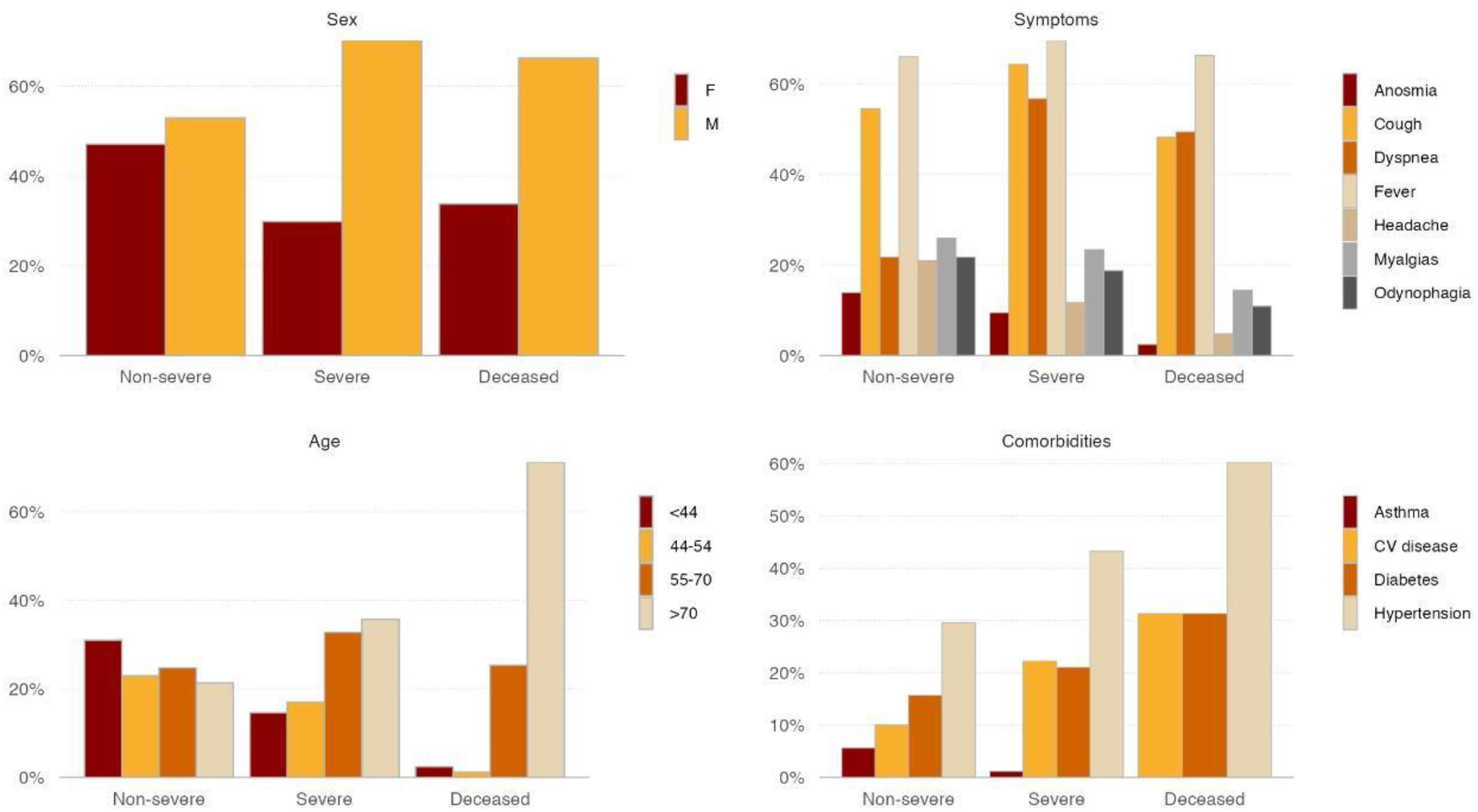
Percentage frequency distribution by sex, age, symptoms and comorbidities clasified by severity.

### RADIOLOGIC FINDINGS

Many patients had normal chest X-Ray (48.8%) and normal computed tomographic (CT) scan of the chest was seen in 16.8%. Ground glass opacities were observed in 24.2% of the patients on radiographs and in 61.1% on chest CT. Of those who had normal X-Ray and a CT was performed, 58.6% had also normal CT while the rest had multilobular consolidation or diffuse alveolar pattern. The radiological pattern in non-severe patients was normal in most of the patients (57.3%) and in the CT scans it was normal in 21.1% and 54% of ground glass opacities. In severe patients, the most frequent X-Ray pattern was ground glass opacity in 52% and normal in 22%, and the most frequent tomographic pattern was diffuse alveolar consolidation in 79.1%. 13.3% of the deceased had a normal x-ray or 7.4% had normal tomography on admission.

### LABORATORY FINDINGS

The most frequent laboratory alteration was high CRP with 68.1%, greater than 10 with a median of 23, followed by lymphocytopenia, lower than 1500 in 58.2% of patients and a median of 1336 and ferritin higher than 500 in 51.5% (median 421). LDH higher than 400 was found in 39.6% of patients with a median of 340, erythrocyte sedimentation greater than 50 in 34.1% with a median of 37 and platelets less than 150000 in 16.2% of cases. In severe patients, a higher frequency of thrombocytopenia were found (19%) than in non-severe, which was 15.2%, leukocytosis was present in 29.8% vs. 8.8% in non-severe, lymphocytopenia less than 1500 in 66.7% of severe patients and 55.3% of non severe. The median CRP was 61 in severe patients with 90.4% higher than 10 and median of 17 in non-severe 61.2% higher than 10, median ferritin was 1128 in severe with 77.8% higher than 500 and 362 in non-severe, 39.2% higher than 500. Renal impairment was observed in 11% of non-severe and 17.9 of severe. No patient required haemodialysis. Transaminase elevation was seen in 31.1% of cases. 67.6% of severe patients had an elevation of lactic dehydrogenase higher than 400 versus 39.2% of non-severe.

**Figure 2.**
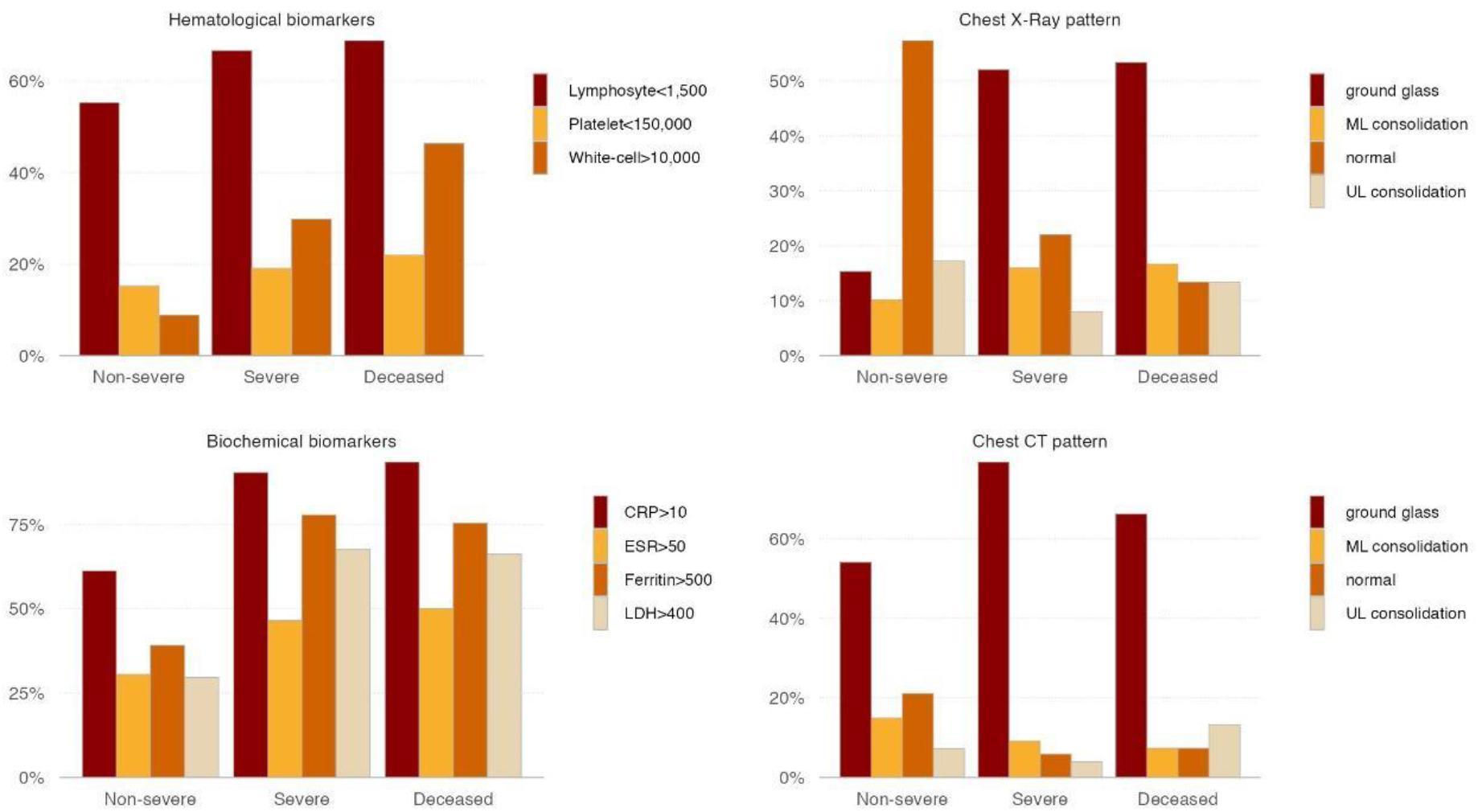
Percentage frequency of hematologic and biochemical alterations and Chest X-Ray and Chest CT pattern clasified by severity.

### CLINICAL OUTCOMES

In our study, 171 hospitalized patients had severe disease (25.5%), 49 patients required assisted ventilation (7.3%), and 83 (12.4%) died. Time from symptom onset to admission was 5 days in non-severe and severe patients and 6 days in deceased patients. The time from start of symptoms to severe disease was an average of 8 days, 11 days from symptom start to assisted ventilation, 3 days from admission to severe disease, and 5 days from admission to IMV. 45% of severe patients were admitted to intensive care units, 88.9% had saturation less than 93 and 36.3% respiratory rate more than 30. No patient required haemodialysis.

Patients who required assisted ventilation had an average length of stay of 50 days and 30 died (61.2%). The average time from symptoms start to death in ventilated patients was 61 days. 42.9% of patients were ventilated in a prone position and 34.7% of ventilated required inotropic treatment. We analysed outcomes in public and private institutions (Supplementary material Table 2-Cases classified into private and public hospital admission). Mortality was 19.3% in the public hospital and 5.8% in private hospitals. The population attending is quite different. Patients of the public hospital had mean age 61, 1.75 comorbidities, with 27.3% of patients without comorbidities, while those in private hospitals had mean age 50, 0,94 simultaneous comorbidities with 52.6% without comorbidities. Patients in private hospitals were admitted 4 days after initiating symptoms, while those in the public hospital were admitted 5 days after symptoms started.

In our study, 10 patients had probably acquired COVID while hospitalized as they initiated symptoms 10 days after admission and the mortality of these patients was 40% with 3 comorbidities

**Figure3.**
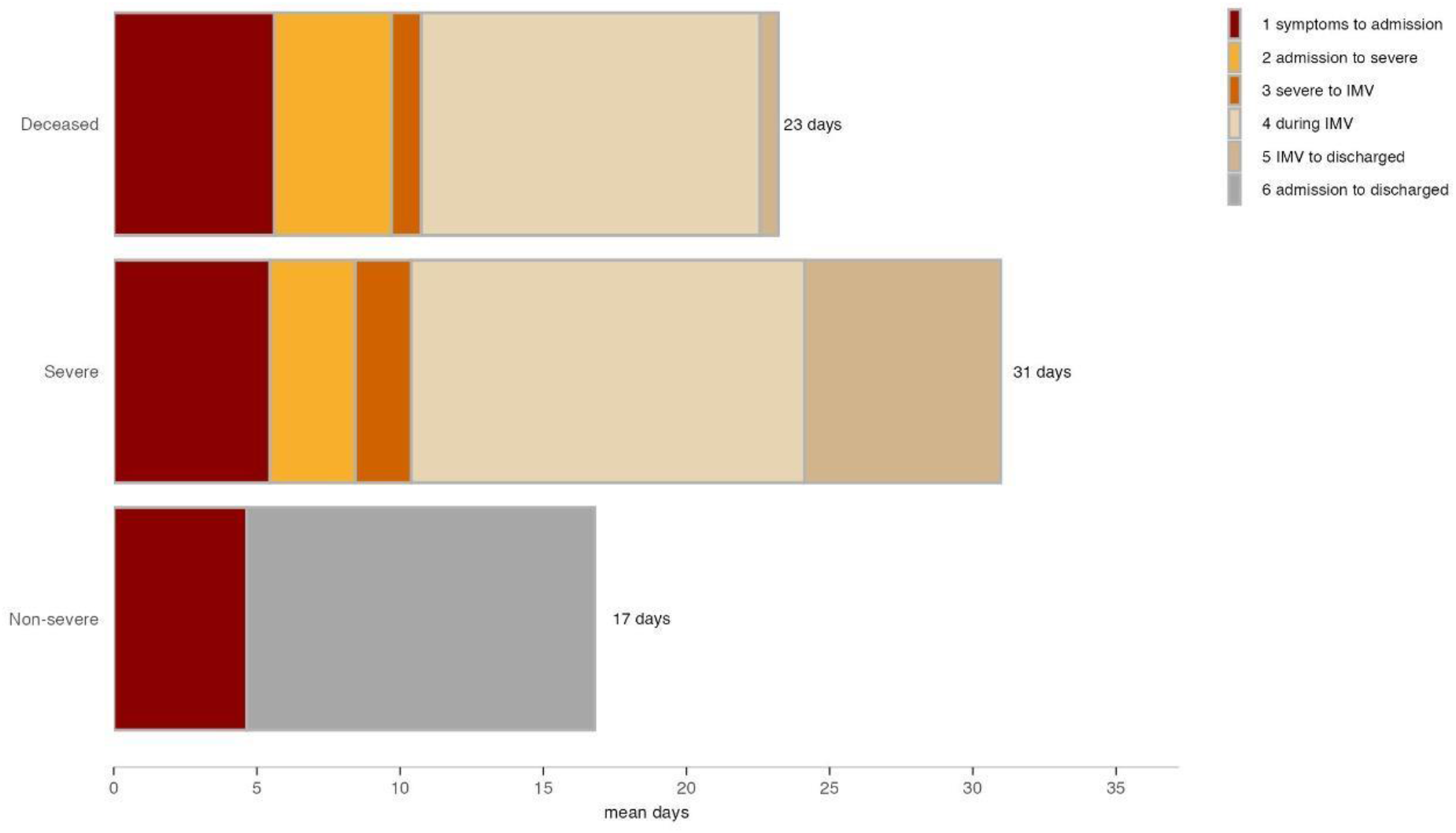
Timeline of illness onset, admission and outcome of patients classified according to severity.

### TREATMENTS

11 severe patients were treated with hydroxychloroquine, 54.5% of whom required assisted ventilation and 18.2% died. Of the 14 severe patients on lopinavir-ritonavir treatment, 50% required mechanical ventilation and 28.6% died vs. 26.9% of the patients who did not and 33.1% died. 66.1% of severe patients were treated with azithromycin, 28.3% of whom died versus 41.4% who did not receive. % (120) of severe patients received steroids, 30.8% of whom died versus 37.3% of the 51 patients that did not receive. 42 (24.6%) of severe patients were treated with plasma and 31% of those required mechanical ventilation and 16.7% died versus patients without plasma of whom 27.9% required ventilation and 38% died. 2 patients had probable severe reactions to plasma, one had heart failure and the other one deceased during plasma treatment.

## Discussion

In our study, as in others, COVID-19 was more frequent in male patients with progressive predominance in severe and dead patients as reported in studies of New York and China^1,3^. The median age (54) was similar to other studies in China (47^5^) and USA with older patients with more severe disease. Severe patients seem to have more symptoms of lower respiratory infection such as dyspnea, while non-severe patients had more upper respiratory symptoms such as anosmia and odynophagia. Only two patients who died had anosmia (2.4%), probably in accordance with a study in Spain that associates anosmia with a good prognosis^7^. 15.1% of patients had only one symptom and 33.1% did not have fever, showing the error of excluding patients for testing without fever or with only one symptom, as happened in our country in the first months of the pandemia.

As described in other studies, most COVID-19 patients were no smokers, with more frequent comorbidities being arterial hypertension, diabetes, and cardiovascular disease. Diabetes, although a frequent comorbidity with 21.1% in severe cases, did not show the frequency shown in other studies as the one conducted in Seattle^6^ (58%). 8.4% of patients were health care workers which represent a higher proportion than observed in other studies (Wuhan 3.5%^3^). Although most plain lung radiographies were normal in non-severe patients, and a fraction of severe patients also had normal radiography, only 13.3% of patients who died had normal radiography on admission. Diffuse alveolar pattern was the most frequent tomographic pattern, as reported in China study^3^, with a normal pattern in a few patients with only 7.4% normal tomography in patients who died. Laboratory abnormalities included lymphopenia, thrombocytopenia, and leukocytosis, although lymphopenia was the more frequent abnormality, leukocytosis frequency was markedly increased in severe patients. CRP value was highly related to the severity of the disease. Lymphocytopenia (58.2%) was not as frequent as in others studies (83.2% in Wuhan^3^). There were not high rates of acute kidney injury, nor haemodialysis as observed in New York studies^2, 4^.

We must mention that in the first months of the pandemia, patients were admitted to hospitals to ensure isolation, so the admitted patients had milder disease, especially in private hospitals, as patients attending to public hospitals were sometimes isolated in centres of isolation.

## Conclusions

The findings of our study support that when considering the diagnosis of COVID-19, the absence of fever and normal radiography, tomography, and laboratory by no means excludes the disease. When considering prognosis, dyspnoea, diffuse alveolar pattern, leukocytosis, and higher CRP were associated with poor outcome, while anosmia, odynophagia and myalgia may be associated with better outcome.

Patients from the public hospital had higher mortality than patients from private hospitals, probably associated with older age, later admission, more severe patients (milder cases were isolated in centres of isolation), and more comorbidities. Low socio-economic status increases the risk of cardiovascular disease, obesity, diabetes, and hypertension as described in previous studies ^8^.

Limitations: As this is an observational study, no statistical significant conclusion can be made as the study design is not adequate for that. As patients could not be weighed during their hospitalization, obesity was merely registered as observation by the attendant physician, so it could have been underrated. On behalf of treatments, we observed that many treatments such as convalescent plasma were offered to younger patients with fewer comorbidities, so the outcome was biased.

## Data Availability

All Data available at link figshare

https://doi.org/10.6084/m9.figshare.15062484.v2

## Data Availability

The data used to support the findings of this study are included within the article. Data are also available from the corresponding author upon reasonable request

## Acknowledgement

We thank all physicians who provided and cared for the study patients and contributed in collecting data: E. Angelo Aliaga Palacios, Margarita Barris, María Belén Bertoli, Barbara Broese, Daniela Brito, Jorge Buccella, Jimena Carrizo, Claudia Condori, Maria Pía Contarbio, Luis Coo, Adriana Corigliano, Sabrina Garce, Carolina Escobar, Roberto Gavazzi, Angeles Gonzalez, Anabella Gottas, Victor Grodek, Alberth Jurado, Leonel Langelotti, Juan Martín Linares, Mariela Mansur, Alejandro Marquez, Daniela Mingoni, Catalina Ramirez Toro, María Estela Ramírez Martínez, Nadia Rodríguez, Clarisa Taffarel, Ana Terusi, Lucila Torasso, María Emilia Ramos Vazquez, Hernando Chacon Corena and we thank Enrique Rodriguez who critically reviewed the study proposal.

We thank Jorge Walpen for his dedicated pro bonus to statistical analysis and Joaquin Sackmann Sala for his contribution to initial data management. Thanks to the patients who consented to donate their data for analysis

## References

1. Richardson S, Hirsch JS, Narasimhan M, et al. Presenting Characteristics, Comorbidities, and Outcomes Among 5700 Patients Hospitalized With COVID-19 in the New York City Area. JAMA. 2020;323(20):2052–2059. doi:10.1001/jama.2020.6775

2. Cummings MJ et al. Epidemiology, clinical course, and outcomes of critically ill adults with COVID-19 in New York City: A prospective cohort study. Lancet 2020 Jun 6; 395:1763.

3. Guan WJ, Ni ZY, Hu Y, Liang WH, Ou CQ, He JX, et al. Clinical characteristics of coronavirus disease 2019 in China. N Engl J Med. 2020.

4. Argenziano MG, Bruce SL, Slater CL, et al. Characterization and clinical course of 1000 patients with coronavirus disease 2019 in New York: retrospective case series. BMJ. 2020;369:m1996.

5. Guan W, Ni Z, Hu Y. Clinical characteristics of coronavirus disease 2019 in China. N Engl J Med. 2020 doi: 10.1056/NEJMoa2002032. published online Feb 28.

6. Bhatraju PK, Ghassemieh BJ, Nichols M, et al. Covid-19 in critically ill patients in the Seattle region – case series. N Engl J Med. 2020; March 30

7. Rubio-Rivas M, Corbella X, Mora-Luján J.M, Loureiro-Amigo J, López Sampalo A, Yera Bergua C, et al. Predicting Clinical Outcome with Phenotypic Clusters in COVID-19 Pneumonia: An Analysis of 12,066 Hospitalized Patients from the Spanish Registry SEMI-COVID-19 J. Clin. Med. 2020, 9, 3488

8. Marmot M., Allen J., Goldblatt P., Boyce T., McNeish D., Grady M. 2010. Fair Society, Healthy lives - the Marmot Review

