## Supplemental Tables 1 and 2 for "COVID-19 in hospitalized patients in 4 hospitals in San Isidro, Buenos Aires, Argentina"

### Tabla 1. Cases classified by hospital

| Characteristic | H.C. San Isidro | S. Las Lomas | S. San Lucas | S. Trinidad | Total |
|---|---|---|---|---|---|
| Cases [num. (%)] | 326 (100) | 110 (100) | 77 (100) | 155 (100) | 668 (100) |
| First ocurrences | … | … | … | … | … |
| …Symptoms | 19/03/2020 | 14/03/2020 | 29/02/2020 | 12/03/2020 | 29/02/2020 |
| …Admission | 08/11/2019 | 14/03/2020 | 05/03/2020 | 16/03/2020 | 08/11/2019 |
| …Positive test | 30/03/2020 | 19/03/2020 | 12/03/2020 | 20/03/2020 | 12/03/2020 |
| Residence [num. (%)] | Residence [num. (%)] | Residence [num. (%)] | Residence [num. (%)] | Residence [num. (%)] | Residence [num. (%)] |
| …San Isidro district | 317 (97.2) | 33 (30.0) | 25 (32.5) | 4 (2.6) | 379 (56.7) |
| …Other | 9 (2.8) | 77 (70.0) | 52 (67.5) | 151 (97.4) | 289 (43.3) |
| Last condition [num. (%)] | … | … | … | … | … |
| …Sick | 240 (73.6) | 104 (94.5) | 71 (92.2) | 128 (82.6) | 543 (81.3) |
| …Deceased | 63 (19.3) | 6 (5.5) | 0 (0.0) | 14 (9.0) | 83 (12.4) |
| …Recovered | 23 (7.1) | 0 (0.0) | 6 (7.8) | 13 (8.4) | 42 (6.3) |
| …Suspected | 0 (0.0) | 0 (0.0) | 0 (0.0) | 0 (0.0) | 0 (0.0) |
| Intervals in days [mean (range)] | … | … | … | … | … |
| …A: symptoms to IMV | 11 (0-106) | 7 (1-16) | 13 (13-13) | 12 (4-29) | 11 (0-106) |
| …B: during IMV | 12 (0-40) | 14 (13-16) | – | 21 (4-63) | 14 (0-63) |
| …C: admission to IMV | 4 (0-16) | 3 (0-6) | 11 (11-11) | 7 (0-25) | 5 (0-25) |
| …D: admission to severe | 3 (0-31) | 2 (0-9) | 4 (2-8) | 3 (0-11) | 3 (0-31) |
| …E: sick | 84 (0-199) | 119 (5-225) | 118 (7-227) | 96 (1-209) | 96 (0-227) |
| …G: severe | 20 (0-161) | 15 (0-221) | 3 (0-117) | 21 (0-120) | 17 (0-221) |
| …H: symptoms to severe | 8 (0-105) | 6 (0-19) | 6 (4-9) | 7 (2-17) | 8 (0-105) |
| …I: in admission | 86 (1-266) | 118 (5-230) | 119 (12-229) | 97 (4-213) | 97 (1-266) |
| …J: symptoms to addmission | 5 (0-104) | 5 (0-37) | 3 (0-12) | 5 (0-36) | 5 (0-104) |
| …K: admission to symptoms | 15 (1-135) | 1 (1-1) | 2 (2-2) | 4 (1-6) | 13 (1-135) |
| …M: deceased | 19 (0-191) | 5 (0-113) | 0 (0-0) | 8 (0-209) | 12 (0-209) |
| …P: severe to IMV | 0 (0-9) | 0 (0-5) | 0 (0-9) | 0 (0-24) | 0 (0-24) |
| …Q: end of IMV to discharged | 1 (0-95) | 0 (0-11) | 0 (0-0) | 0 (0-14) | 0 (0-95) |
| …R: recovered | 11 (0-210) | 0 (0-0) | 16 (0-225) | 16 (0-212) | 11 (0-225) |
| …S: suspected | 5 (0-105) | 3 (0-59) | 3 (0-12) | 5 (0-51) | 5 (0-105) |
| …U: symptoms to hidrox. | 12 (6-20) | – | – | 9 (5-17) | 10 (5-20) |
| …V: symptoms to antivirals | 8 (5-11) | – | 4 (4-4) | 9 (6-17) | 8 (4-17) |
| …W: symptoms to plasma | 13 (1-106) | – | 7 (7-7) | 6 (3-15) | 12 (1-106) |
| …Z: non severe in admission | 10 (0-266) | 7 (0-29) | 10 (0-36) | 7 (0-38) | 9 (0-266) |
| Age | … | … | … | … | … |
| …median [IRQ] | 61 (48-75) | 54 (44-67) | 43 (33-53) | 51 (40-64) | 54 (43-70) |
| …range [min-mean-max] | 15-60-95 | 19-57-98 | 22-45-76 | 16-53-98 | 15-56-98 |
| …Quantiles [num. (%)] | … | … | … | … | … |
| ……<44 | 66 (20.2) | 22 (20.0) | 39 (50.6) | 52 (33.5) | 179 (26.8) |
| ……44-54 | 56 (17.2) | 36 (32.7) | 22 (28.6) | 42 (27.1) | 156 (23.4) |
| ……55-70 | 88 (27.0) | 33 (30.0) | 12 (15.6) | 33 (21.3) | 166 (24.9) |
| ……>70 | 116 (35.6) | 19 (17.3) | 4 (5.2) | 28 (18.1) | 167 (25.0) |
| …10-year age intervals [num. (%)] | … | … | … | … | … |
| ……0-10 | 0 (0.0) | 0 (0.0) | 0 (0.0) | 0 (0.0) | 0 (0.0) |
| ……10-20 | 6 (1.8) | 1 (0.9) | 0 (0.0) | 1 (0.6) | 8 (1.2) |
| ……20-30 | 21 (6.4) | 3 (2.7) | 8 (10.4) | 18 (11.6) | 50 (7.5) |
| ……30-40 | 22 (6.7) | 8 (7.3) | 22 (28.6) | 17 (11.0) | 69 (10.3) |
| ……40-50 | 45 (13.8) | 24 (21.8) | 20 (26.0) | 38 (24.5) | 127 (19.0) |
| ……50-60 | 53 (16.3) | 33 (30.0) | 15 (19.5) | 30 (19.4) | 131 (19.6) |
| ……60-70 | 57 (17.5) | 19 (17.3) | 8 (10.4) | 21 (13.5) | 105 (15.7) |
| ……70-80 | 68 (20.9) | 8 (7.3) | 4 (5.2) | 14 (9.0) | 94 (14.1) |
| ……80-90 | 45 (13.8) | 9 (8.2) | 0 (0.0) | 11 (7.1) | 65 (9.7) |
| ……90-100 | 9 (2.8) | 5 (4.5) | 0 (0.0) | 5 (3.2) | 19 (2.8) |

| | | | | | |
|---|---|---|---|---|---|
| …Female sex [num. (%)] | 134 (41.1) | 50 (45.5) | 38 (49.4) | 63 (40.6) | 285 (42.7) |
| Fever on admission [num. (%)] | … | … | … | … | … |
| …fever detected | 194 (59.5) | 90 (81.8) | 51 (66.2) | 112 (72.3) | 447 (66.9) |
| ……Median temperature [ºC (IQR)] | 38 (38-38) | 38 (38-38) | 38 (38-38) | 38 (38-38) | 38 (38-38) |
| ……Ranges [min-max] | 30-40 | 37.4-40 | 36.5-39 | 37.5-39.2 | 30-40 |
| …no fever detected | 125 (38.3) | 18 (16.4) | 21 (27.3) | 43 (27.7) | 207 (31.0) |
| …not specified | 0 (0.0) | 0 (0.0) | 0 (0.0) | 0 (0.0) | 0 (0.0) |
| Symptoms [num. (%)] | … | … | … | … | … |
| …Cough | 177 (54.3) | 53 (48.2) | 44 (57.1) | 107 (69.0) | 381 (57.0) |
| …Myalgia | 60 (18.4) | 28 (25.5) | 34 (44.2) | 47 (30.3) | 169 (25.3) |
| …Headache | 42 (12.9) | 16 (14.5) | 27 (35.1) | 39 (25.2) | 124 (18.6) |
| …Odynophagia | 53 (16.3) | 18 (16.4) | 28 (36.4) | 41 (26.5) | 140 (21.0) |
| …Rhinorrhoea | 3 (0.9) | 7 (6.4) | 7 (9.1) | 3 (1.9) | 20 (3.0) |
| …Diarrhoea | 14 (4.3) | 11 (10.0) | 7 (9.1) | 18 (11.6) | 50 (7.5) |
| …Dyspnoea | 117 (35.9) | 37 (33.6) | 7 (9.1) | 44 (28.4) | 205 (30.7) |
| …Anosmia | 30 (9.2) | 10 (9.1) | 17 (22.1) | 28 (18.1) | 85 (12.7) |
| …Fatigue | 34 (10.4) | 11 (10.0) | 1 (1.3) | 0 (0.0) | 46 (6.9) |
| …Malaise | 18 (5.5) | 0 (0.0) | 8 (10.4) | 0 (0.0) | 26 (3.9) |
| …Vomiting | 3 (0.9) | 1 (0.9) | 2 (2.6) | 2 (1.3) | 8 (1.2) |
| …Dysgeusia | 17 (5.2) | 8 (7.3) | 10 (13.0) | 10 (6.5) | 45 (6.7) |
| …Simultaneous symptoms [mean] | 2.34 (0.7) | 2.64 (2.4) | 3.16 (4.1) | 2.91 (1.9) | 2.61 (0.4) |
| …One sympton only | 60 (18.4) | 17 (15.5) | 7 (9.1) | 17 (11.0) | 101 (15.1) |
| Comorbidities [num. (%)] | … | … | … | … | … |
| …DBT | 73 (22.4) | 15 (13.6) | 3 (3.9) | 23 (14.8) | 114 (17.1) |
| …HTA | 138 (42.3) | 33 (30.0) | 14 (18.2) | 36 (23.2) | 221 (33.1) |
| …IECA | 56 (17.2) | 6 (5.5) | 5 (6.5) | 7 (4.5) | 74 (11.1) |
| …ARB | 43 (13.2) | 20 (18.2) | 7 (9.1) | 14 (9.0) | 84 (12.6) |
| …Chronic kidney disease | 25 (7.7) | 0 (0.0) | 0 (0.0) | 5 (3.2) | 30 (4.5) |
| …COPD | 9 (2.8) | 5 (4.5) | 0 (0.0) | 4 (2.6) | 18 (2.7) |
| …Asthma | 17 (5.2) | 2 (1.8) | 5 (6.5) | 6 (3.9) | 30 (4.5) |
| …Cardiovascular disease | 64 (19.6) | 7 (6.4) | 3 (3.9) | 14 (9.0) | 88 (13.2) |
| …Lung disease | 10 (3.1) | 5 (4.5) | 1 (1.3) | 4 (2.6) | 20 (3.0) |
| …Cancer | 15 (4.6) | 3 (2.7) | 0 (0.0) | 8 (5.2) | 26 (3.9) |
| …Current smoker | 15 (4.6) | 4 (3.6) | 10 (13.0) | 3 (1.9) | 32 (4.8) |
| …former smoker | 71 (21.8) | 1 (0.9) | 9 (11.7) | 8 (5.2) | 89 (13.3) |
| …Never smoked | 234 (71.8) | 90 (81.8) | 52 (67.5) | 142 (91.6) | 518 (77.5) |
| …Chronic liver disease | 5 (1.5) | 1 (0.9) | 0 (0.0) | 0 (0.0) | 6 (0.9) |
| …Immunosupresive drugs | 13 (4.0) | 4 (3.6) | 1 (1.3) | 7 (4.5) | 25 (3.7) |
| …Pregnancy | 6 (1.8) | 0 (0.0) | 2 (2.6) | 2 (1.3) | 10 (1.5) |
| …HIV | 3 (0.9) | 0 (0.0) | 0 (0.0) | 0 (0.0) | 3 (0.4) |
| …Obesity | 6 (1.8) | 0 (0.0) | 0 (0.0) | 15 (9.7) | 21 (3.1) |
| …Simultaneous comorbidities [mean] | 1.75 | 0.96 | 0.78 | 1.01 | 1.33 |
| …Whithout comorbidities [num. (%)] | 89 (27.3) | 62 (56.4) | 42 (54.5) | 76 (49.0) | 269 (40.3) |
| …Whithout comorbidities younger than 70 | 64 (19.6) | 54 (49.1) | 40 (51.9) | 69 (44.5) | 227 (34.0) |
| Epidemiology [num. (%)] | … | … | … | … | … |
| …Travel | 0 (0.0) | 2 (1.8) | 6 (7.8) | 6 (3.9) | 14 (2.1) |
| …Direct contact | 76 (23.3) | 35 (31.8) | 20 (26.0) | 43 (27.7) | 174 (26.0) |
| …Health personnel | 7 (2.1) | 9 (8.2) | 19 (24.7) | 21 (13.5) | 56 (8.4) |
| Radiologic findings [num. (%)] | … | … | … | … | … |
| …Chest X-Ray 1 | 158 (100.0) | 3 (100.0) | 11 (100.0) | 35 (100.0) | 207 (100.0) |
| ……1. normal | 63 (39.9) | 1 (33.3) | 9 (81.8) | 28 (80.0) | 101 (48.8) |
| ……2. alveolar ground-glass | 27 (17.1) | 1 (33.3) | 0 (0.0) | 3 (8.6) | 31 (15.0) |
| ……3. multifocal | 22 (13.9) | 0 (0.0) | 0 (0.0) | 2 (5.7) | 24 (11.6) |
| ……4. ground-glass opacities | 45 (28.5) | 1 (33.3) | 2 (18.2) | 2 (5.7) | 50 (24.2) |
| ……5. other | 1 (0.6) | 0 (0.0) | 0 (0.0) | 0 (0.0) | 1 (0.5) |
| …Chest X-Ray 2 | 11 (100.0) | 2 (100.0) | 4 (100.0) | 7 (100.0) | 24 (100.0) |

| | | | | | |
|---|---|---|---|---|---|
| ......1. normal | 0 (0.0) | 0 (0.0) | 0 (0.0) | 2 (28.6) | 2 (8.3) |
| ......2. alveolar segmento | 0 (0.0) | 0 (0.0) | 0 (0.0) | 0 (0.0) | 0 (0.0) |
| ......3. multifocal | 0 (0.0) | 1 (50.0) | 0 (0.0) | 0 (0.0) | 1 (4.2) |
| ......4. ground-glass opacities | 10 (90.9) | 1 (50.0) | 2 (50.0) | 5 (71.4) | 18 (75.0) |
| ......5. other | 1 (9.1) | 0 (0.0) | 2 (50.0) | 0 (0.0) | 3 (12.5) |
| ...Chest CT 1 | 241 (100.0) | 108 (100.0) | 64 (100.0) | 129 (100.0) | 542 (100.0) |
| ......1. normal | 35 (14.5) | 14 (13.0) | 22 (34.4) | 20 (15.5) | 91 (16.8) |
| ......2. alveolar segmento | 15 (6.2) | 9 (8.3) | 0 (0.0) | 10 (7.8) | 34 (6.3) |
| ......3. multifocal | 19 (7.9) | 2 (1.9) | 7 (10.9) | 44 (34.1) | 72 (13.3) |
| ......4. ground-glass opacities | 167 (69.3) | 77 (71.3) | 35 (54.7) | 52 (40.3) | 331 (61.1) |
| ......5. other | 5 (2.1) | 6 (5.6) | 0 (0.0) | 3 (2.3) | 14 (2.6) |
| ...Chest CT 2 | 43 (100.0) | 3 (100.0) | 1 (100.0) | 8 (100.0) | 55 (100.0) |
| ......1. normal | 1 (2.3) | 0 (0.0) | 0 (0.0) | 1 (12.5) | 2 (3.6) |
| ......2. alveolar segmento | 0 (0.0) | 0 (0.0) | 0 (0.0) | 0 (0.0) | 0 (0.0) |
| ......3. multifocal | 0 (0.0) | 0 (0.0) | 0 (0.0) | 1 (12.5) | 1 (1.8) |
| ......4. ground-glass opacities | 41 (95.3) | 3 (100.0) | 1 (100.0) | 6 (75.0) | 51 (92.7) |
| ......5. other | 1 (2.3) | 0 (0.0) | 0 (0.0) | 0 (0.0) | 1 (1.8) |
| ...Chest X-Ray 1 | ... | ... | ... | ... | ... |
| ......plus chest CT 1 | 8 (12.7) | 0 (0.0) | 3 (33.3) | 6 (21.4) | 17 (16.8) |
| ......plus chest CT 2 | 0 (0.0) | 0 (0.0) | 0 (0.0) | 0 (0.0) | 0 (0.0) |
| ......plus chest CT 3 | 3 (4.8) | 0 (0.0) | 0 (0.0) | 0 (0.0) | 3 (3.0) |
| ......plus chest CT 4 | 7 (11.1) | 0 (0.0) | 2 (22.2) | 0 (0.0) | 9 (8.9) |
| ......plus chest CT 5 | 0 (0.0) | 0 (0.0) | 0 (0.0) | 0 (0.0) | 0 (0.0) |
| Laboratory results [num. (%)] | ... | ... | ... | ... | ... |
| ...... [median (IQR)] | ... | ... | ... | ... | ... |
| ...Lymphocyte count | 323 (100.0) | 110 (100.0) | 68 (100.0) | 109 (100.0) | 610 (100.0) |
| ...... | 1369 (995-1827) | 1140 (837-1620) | 1300 (975-1700) | 1537 (952-1938) | 1336 (936-1796) |
| ......Lymphocyte <1,500 | 184 (57.0) | 80 (72.7) | 40 (58.8) | 51 (46.8) | 355 (58.2) |
| ...White-cell count | 324 (100.0) | 94 (100.0) | 70 (100.0) | 155 (100.0) | 643 (100.0) |
| ...... | 6900 (5100-9625) | 5300 (4400-7200) | 4900 (3925-5900) | 5600 (4300-6700) | 5900 (4600-8000) |
| ...... <4,500 | 48 (14.8) | 26 (27.7) | 25 (35.7) | 44 (28.4) | 143 (22.2) |
| ...... >10,000 | 73 (22.5) | 6 (6.4) | 1 (1.4) | 12 (7.7) | 92 (14.3) |
| ...Platelet | 324 (100.0) | 94 (100.0) | 69 (100.0) | 155 (100.0) | 642 (100.0) |
| ...... | 211800 (164675-278500) | 183000 (146250-231750) | 191000 (173000-228000) | 219000 (175500-264500) | 207050 (166000-257750) |
| ...... <150,000 | 58 (17.9) | 26 (27.7) | 6 (8.7) | 14 (9.0) | 104 (16.2) |
| ...ESR | 206 (100.0) | 80 (100.0) | 54 (100.0) | 29 (100.0) | 369 (100.0) |
| ...... | 47 (27-72) | 28 (16-41) | 20 (10-40) | 36 (15-78) | 37 (20-63) |
| ...... >50 | 93 (45.1) | 15 (18.8) | 7 (13.0) | 11 (37.9) | 126 (34.1) |
| ...Creatinine | 323 (100.0) | 109 (100.0) | 69 (100.0) | 151 (100.0) | 652 (100.0) |
| ...... | 1 (1-1) | 1 (1-1) | 1 (1-1) | 1 (1-1) | 1 (1-1) |
| ...... >1.20 | 53 (16.4) | 12 (11.0) | 2 (2.9) | 16 (10.6) | 83 (12.7) |
| ...Lactate dehydrogenase | 314 (100.0) | 100 (100.0) | 59 (100.0) | 90 (100.0) | 563 (100.0) |
| ...... | 434 (321-650) | 184 (156-236) | 186 (154-222) | 354 (295-461) | 340 (228-532) |
| ...... >400 | 182 (58.0) | 4 (4.0) | 1 (1.7) | 36 (40.0) | 223 (39.6) |
| ...... >600 | 100 (31.8) | 0 (0.0) | 0 (0.0) | 5 (5.6) | 105 (18.7) |
| ...C-reactive protein | 177 (100.0) | 98 (100.0) | 44 (100.0) | 123 (100.0) | 442 (100.0) |
| ...... | 50 (13-113) | 21 (12-62) | 6 (3-16) | 15 (5-41) | 23 (7-73) |
| ...... >10 | 136 (76.8) | 76 (77.6) | 16 (36.4) | 73 (59.3) | 301 (68.1) |
| ...... >60 | 82 (46.3) | 25 (25.5) | 2 (4.5) | 23 (18.7) | 132 (29.9) |
| ...... >100 | 59 (33.3) | 12 (12.2) | 1 (2.3) | 10 (8.1) | 82 (18.6) |
| ...Ferritin | 221 (100.0) | 69 (100.0) | 32 (100.0) | 99 (100.0) | 421 (100.0) |
| ...... | 612 (281-1607) | 342 (197-627) | 277 (125-537) | 651 (258-1423) | 514 (231-1152) |
| ...... >500 | 125 (56.6) | 26 (37.7) | 8 (25.0) | 58 (58.6) | 217 (51.5) |
| ...... >600 | 112 (50.7) | 21 (30.4) | 8 (25.0) | 51 (51.5) | 192 (45.6) |
| ...Bilirrubin | 297 (100.0) | 107 (100.0) | 63 (100.0) | 136 (100.0) | 603 (100.0) |
| ...... | 1 (0-1) | 0 (0-1) | 0 (0-1) | 0 (0-1) | 0 (0-1) |
| ...Alanine aminotransferase | 324 (100.0) | 107 (100.0) | 64 (100.0) | 136 (100.0) | 631 (100.0) |
| ...... | 33 (20-58) | 23 (13-34) | 23 (16-42) | 28 (17-41) | 28 (17-48) |

| | | | | | |
|---|---|---|---|---|---|
| …… >41 | 124 (38.3) | 21 (19.6) | 17 (26.6) | 34 (25.0) | 196 (31.1) |
| …Aspartate aminotransferase | 322 (100.0) | 107 (100.0) | 64 (100.0) | 136 (100.0) | 629 (100.0) |
| <- <- | 34 (23-52) | 23 (18-34) | 25 (21-33) | 26 (20-36) | 29 (21-44) |
| …… >40 | 126 (39.1) | 20 (18.7) | 7 (10.9) | 25 (18.4) | 178 (28.3) |
| …Procalcitonin | 0 (0.0) | 1 (100.0) | 3 (100.0) | 2 (100.0) | 6 (100.0) |
| <- <- | – | 0 (0-0) | 0 (0-0) | 0 (0-0) | 0 (0-0) |
| …D-dimer | 9 (100.0) | 55 (100.0) | 57 (100.0) | 95 (100.0) | 216 (100.0) |
| …… | 1 (1-1) | 0 (0-0) | 0 (0-0) | 1 (0-1) | 0 (0-1) |
| …3 normal biomarkers | 3 (0.9) | 2 (1.8) | 8 (10.4) | 1 (0.6) | 14 (2.1) |
| Treatements [num. (%)] | … | … | … | … | … |
| …With hidroxychloroquine | 5 (1.5) | 1 (0.9) | 2 (2.6) | 10 (6.5) | 18 (2.7) |
| ……With IMV | 1 (20.0) | 1 (100.0) | 1 (50.0) | 3 (30.0) | 6 (33.3) |
| ……deceased | 1 (20.0) | 0 (0.0) | 0 (0.0) | 1 (10.0) | 2 (11.1) |
| …Without hidroxychloroquine | 321 (98.5) | 109 (99.1) | 75 (97.4) | 145 (93.5) | 650 (97.3) |
| ……With IMV | 31 (9.7) | 3 (2.8) | 0 (0.0) | 9 (6.2) | 43 (6.6) |
| ……deceased | 62 (19.3) | 6 (5.5) | 0 (0.0) | 13 (9.0) | 81 (12.5) |
| …With lopinavir/ritonavir | 8 (2.5) | 1 (0.9) | 3 (3.9) | 7 (4.5) | 19 (2.8) |
| ……With IMV | 2 (25.0) | 1 (100.0) | 1 (33.3) | 3 (42.9) | 7 (36.8) |
| ……deceased | 3 (37.5) | 0 (0.0) | 0 (0.0) | 1 (14.3) | 4 (21.1) |
| …Without lopinavir/ritonavir | 318 (97.5) | 109 (99.1) | 74 (96.1) | 148 (95.5) | 649 (97.2) |
| ……With IMV | 30 (9.4) | 3 (2.8) | 0 (0.0) | 9 (6.1) | 42 (6.5) |
| ……deceased | 60 (18.9) | 6 (5.5) | 0 (0.0) | 13 (8.8) | 79 (12.2) |
| …With plasma | 35 (10.7) | 1 (0.9) | 1 (1.3) | 8 (5.2) | 45 (6.7) |
| ……With IMV | 11 (31.4) | 0 (0.0) | 0 (0.0) | 2 (25.0) | 13 (28.9) |
| ……deceased | 6 (17.1) | 0 (0.0) | 0 (0.0) | 1 (12.5) | 7 (15.6) |
| …Without plasma | 291 (89.3) | 109 (99.1) | 76 (98.7) | 147 (94.8) | 623 (93.3) |
| ……With IMV | 21 (7.2) | 4 (3.7) | 1 (1.3) | 10 (6.8) | 36 (5.8) |
| ……deceased | 57 (19.6) | 6 (5.5) | 0 (0.0) | 13 (8.8) | 76 (12.2) |
| …With azithromycin | 225 (69.0) | 79 (71.8) | 37 (48.1) | 42 (27.1) | 383 (57.3) |
| ……With IMV | 22 (9.8) | 4 (5.1) | 0 (0.0) | 6 (14.3) | 32 (8.4) |
| ……deceased | 40 (17.8) | 3 (3.8) | 0 (0.0) | 4 (9.5) | 47 (12.3) |
| …Without azithromycin | 101 (31.0) | 31 (28.2) | 40 (51.9) | 113 (72.9) | 285 (42.7) |
| ……With IMV | 10 (9.9) | 0 (0.0) | 1 (2.5) | 6 (5.3) | 17 (6.0) |
| ……deceased | 23 (22.8) | 3 (9.7) | 0 (0.0) | 10 (8.8) | 36 (12.6) |
| …With steroids | 114 (35.0) | 37 (33.6) | 8 (10.4) | 41 (26.5) | 200 (29.9) |
| ……With IMV | 24 (21.1) | 3 (8.1) | 0 (0.0) | 9 (22.0) | 36 (18.0) |
| ……deceased | 33 (28.9) | 3 (8.1) | 0 (0.0) | 7 (17.1) | 43 (21.5) |
| …Without steroids | 212 (65.0) | 73 (66.4) | 69 (89.6) | 114 (73.5) | 468 (70.1) |
| ……With IMV | 8 (3.8) | 1 (1.4) | 1 (1.4) | 3 (2.6) | 13 (2.8) |
| ……deceased | 30 (14.2) | 3 (4.1) | 0 (0.0) | 7 (6.1) | 40 (8.5) |
| Evolution [num. (%)] | … | … | … | … | … |
| …Temperature >38 | 27 (8.3) | 31 (28.2) | 1 (1.3) | 20 (12.9) | 79 (11.8) |
| …RR >30? | 47 (14.4) | 2 (1.8) | 2 (2.6) | 11 (7.1) | 62 (9.3) |
| <- SAT <93? | 101 (31.0) | 9 (8.2) | 3 (3.9) | 39 (25.2) | 152 (22.8) |
| …IMV | 32 (9.8) | 4 (3.6) | 1 (1.3) | 12 (7.7) | 49 (7.3) |
| …NIV | 0 (0.0) | 1 (0.9) | 2 (2.6) | 0 (0.0) | 3 (0.4) |
| …infiltrados <48 hs | 21 (6.4) | 3 (2.7) | 1 (1.3) | 9 (5.8) | 34 (5.1) |
| …Altered conciousness | 15 (4.6) | 1 (0.9) | 0 (0.0) | 0 (0.0) | 16 (2.4) |
| …Hemodynamic instability | 17 (5.2) | 1 (0.9) | 0 (0.0) | 4 (2.6) | 22 (3.3) |
| …CURB >=2 | 6 (1.8) | 1 (0.9) | 1 (1.3) | 5 (3.2) | 13 (1.9) |
| …ICU admission | 50 (15.3) | 9 (8.2) | 4 (5.2) | 14 (9.0) | 77 (11.5) |
| …IMV prone position | 17 (5.2) | 1 (0.9) | 1 (1.3) | 2 (1.3) | 21 (3.1) |
| …Haemodyalisis | 0 (0.0) | 0 (0.0) | 0 (0.0) | 0 (0.0) | 0 (0.0) |
| …Arrhythmia | 4 (1.2) | 0 (0.0) | 0 (0.0) | 0 (0.0) | 4 (0.6) |
| …Myocarditis/IM | 0 (0.0) | 0 (0.0) | 0 (0.0) | 0 (0.0) | 0 (0.0) |
| …Inotropic agents | 16 (4.9) | 0 (0.0) | 0 (0.0) | 2 (1.3) | 18 (2.7) |

### Tabla 2. Cases classified into private and public hospital admission

| Characteristic | Public | Private | Total |
|---|---|---|---|
| Cases [num. (%)] | 326 (100) | 342 (100) | 668 (100) |
| First ocurrences | ... | ... | ... |
| ...Symptoms | 19/03/2020 | 29/02/2020 | 29/02/2020 |
| ...Admission | 08/11/2019 | 05/03/2020 | 08/11/2019 |
| ...Positive test | 30/03/2020 | 12/03/2020 | 12/03/2020 |
| Residence [num. (%)] | Residence [num. (%)] | Residence [num. (%)] | Residence [num. (%)] |
| ...San Isidro district | 317 (97.2) | 62 (18.1) | 379 (56.7) |
| ...Other | 9 (2.8) | 280 (81.9) | 289 (43.3) |
| Last condition [num. (%)] | ... | ... | ... |
| ...Sick | 240 (73.6) | 303 (88.6) | 543 (81.3) |
| ...Deceased | 63 (19.3) | 20 (5.8) | 83 (12.4) |
| ...Recovered | 23 (7.1) | 19 (5.6) | 42 (6.3) |
| ...Suspected | 0 (0.0) | 0 (0.0) | 0 (0.0) |
| Intervals in days [mean (range)] | ... | ... | ... |
| ...A: symptoms to IMV | 11 (0-106) | 11 (1-29) | 11 (0-106) |
| ...B: during IMV | 12 (0-40) | 20 (4-63) | 14 (0-63) |
| ...C: admission to IMV | 4 (0-16) | 6 (0-25) | 5 (0-25) |
| ...D: admission to severe | 3 (0-31) | 3 (0-11) | 3 (0-31) |
| ...E: sick | 84 (0-199) | 108 (1-227) | 96 (0-227) |
| ...G: severe | 20 (0-161) | 15 (0-221) | 17 (0-221) |
| ...H: symptoms to severe | 8 (0-105) | 7 (0-19) | 8 (0-105) |
| ...I: in admission | 86 (1-266) | 108 (4-230) | 97 (1-266) |
| ...J: symptoms to addmission | 5 (0-104) | 4 (0-37) | 5 (0-104) |
| ...K: admission to symptoms | 15 (1-135) | 3 (1-6) | 13 (1-135) |
| ...M: deceased | 19 (0-191) | 5 (0-209) | 12 (0-209) |
| ...P: severe to IMV | 0 (0-9) | 0 (0-24) | 0 (0-24) |
| ...Q: end of IMV to discharged | 1 (0-95) | 0 (0-14) | 0 (0-95) |
| ...R: recovered | 11 (0-210) | 11 (0-225) | 11 (0-225) |
| ...S: suspected | 5 (0-105) | 4 (0-59) | 5 (0-105) |
| ...U: symptoms to hidrox. | 12 (6-20) | 9 (5-17) | 10 (5-20) |
| ...V: symptoms to antivirals | 8 (5-11) | 9 (4-17) | 8 (4-17) |
| ...W: symptoms to plasma | 13 (1-106) | 7 (3-15) | 12 (1-106) |
| ...Z: non severe in admission | 10 (0-266) | 8 (0-38) | 9 (0-266) |
| Age | ... | ... | ... |
| ...median [IRQ] | 61 (48-75) | 50 (40-63) | 54 (43-70) |
| ...range [min-mean-max] | 15-60-95 | 16-52-98 | 15-56-98 |
| ...Quantiles [num. (%)] | ... | ... | ... |
| ......<44 | 66 (20.2) | 113 (33.0) | 179 (26.8) |
| ......44-54 | 56 (17.2) | 100 (29.2) | 156 (23.4) |
| ......55-70 | 88 (27.0) | 78 (22.8) | 166 (24.9) |
| ......>70 | 116 (35.6) | 51 (14.9) | 167 (25.0) |
| ...10-year age intervals [num. (%)] | ... | ... | ... |
| ......0-10 | 0 (0.0) | 0 (0.0) | 0 (0.0) |
| ......10-20 | 6 (1.8) | 2 (0.6) | 8 (1.2) |
| ......20-30 | 21 (6.4) | 29 (8.5) | 50 (7.5) |
| ......30-40 | 22 (6.7) | 47 (13.7) | 69 (10.3) |
| ......40-50 | 45 (13.8) | 82 (24.0) | 127 (19.0) |
| ......50-60 | 53 (16.3) | 78 (22.8) | 131 (19.6) |
| ......60-70 | 57 (17.5) | 48 (14.0) | 105 (15.7) |
| ......70-80 | 68 (20.9) | 26 (7.6) | 94 (14.1) |
| ......80-90 | 45 (13.8) | 20 (5.8) | 65 (9.7) |
| ......90-100 | 9 (2.8) | 10 (2.9) | 19 (2.8) |
| ...Female sex [num. (%)] | 134 (41.1) | 151 (44.2) | 285 (42.7) |

| | | | |
|---|---|---|---|
| Fever on admission [num. (%)] | … | … | … |
| …fever detected | 194 (59.5) | 253 (74.0) | 447 (66.9) |
| ……Median temperature [ºC (IQR)] | 38 (38-38) | 38 (38-38) | 38 (38-38) |
| ……Ranges [min-max] | 30-40 | 36.5-40 | 30-40 |
| …no fever detected | 125 (38.3) | 82 (24.0) | 207 (31.0) |
| …not specified | 0 (0.0) | 0 (0.0) | 0 (0.0) |
| Symptoms [num. (%)] | … | … | … |
| …Cough | 177 (54.3) | 204 (59.6) | 381 (57.0) |
| …Myalgia | 60 (18.4) | 109 (31.9) | 169 (25.3) |
| …Headache | 42 (12.9) | 82 (24.0) | 124 (18.6) |
| …Odynophagia | 53 (16.3) | 87 (25.4) | 140 (21.0) |
| …Rhinorrhoea | 3 (0.9) | 17 (5.0) | 20 (3.0) |
| …Diarrhoea | 14 (4.3) | 36 (10.5) | 50 (7.5) |
| …Dyspnoea | 117 (35.9) | 88 (25.7) | 205 (30.7) |
| …Anosmia | 30 (9.2) | 55 (16.1) | 85 (12.7) |
| …Fatigue | 34 (10.4) | 12 (3.5) | 46 (6.9) |
| …Malaise | 18 (5.5) | 8 (2.3) | 26 (3.9) |
| …Vomiting | 3 (0.9) | 5 (1.5) | 8 (1.2) |
| …Dysgeusia | 17 (5.2) | 28 (8.2) | 45 (6.7) |
| …Simultaneous symptoms [mean] | 2.34 (0.7) | 2.88 (0.8) | 2.61 (0.4) |
| …One sympton only | 60 (18.4) | 41 (12.0) | 101 (15.1) |
| Comorbidities [num. (%)] | … | … | … |
| …DBT | 73 (22.4) | 41 (12.0) | 114 (17.1) |
| …HTA | 138 (42.3) | 83 (24.3) | 221 (33.1) |
| …IECA | 56 (17.2) | 18 (5.3) | 74 (11.1) |
| …ARB | 43 (13.2) | 41 (12.0) | 84 (12.6) |
| …Chronic kidney disease | 25 (7.7) | 5 (1.5) | 30 (4.5) |
| …COPD | 9 (2.8) | 9 (2.6) | 18 (2.7) |
| …Asthma | 17 (5.2) | 13 (3.8) | 30 (4.5) |
| …Cardiovascular disease | 64 (19.6) | 24 (7.0) | 88 (13.2) |
| …Lung disease | 10 (3.1) | 10 (2.9) | 20 (3.0) |
| …Cancer | 15 (4.6) | 11 (3.2) | 26 (3.9) |
| …Current smoker | 15 (4.6) | 17 (5.0) | 32 (4.8) |
| …former smoker | 71 (21.8) | 18 (5.3) | 89 (13.3) |
| …Never smoked | 234 (71.8) | 284 (83.0) | 518 (77.5) |
| …Chronic liver disease | 5 (1.5) | 1 (0.3) | 6 (0.9) |
| …Immunosupresive drugs | 13 (4.0) | 12 (3.5) | 25 (3.7) |
| …Pregnancy | 6 (1.8) | 4 (1.2) | 10 (1.5) |
| …HIV | 3 (0.9) | 0 (0.0) | 3 (0.4) |
| …Obesity | 6 (1.8) | 15 (4.4) | 21 (3.1) |
| …Simultaneous comorbidities [mean] | 1.75 | 0.94 | 1.33 |
| …Whithout comorbidities [num. (%)] | 89 (27.3) | 180 (52.6) | 269 (40.3) |
| …Whithout comorbidities younger than 70 | 64 (19.6) | 163 (47.7) | 227 (34.0) |
| Epidemiology [num. (%)] | … | … | … |
| …Travel | 0 (0.0) | 14 (4.1) | 14 (2.1) |
| …Direct contact | 76 (23.3) | 98 (28.7) | 174 (26.0) |
| …Health personnel | 7 (2.1) | 49 (14.3) | 56 (8.4) |
| Radiologic findings [num. (%)] | … | … | … |
| …Chest X-Ray 1 | 158 (100.0) | 49 (100.0) | 207 (100.0) |
| ……1. normal | 63 (39.9) | 38 (77.6) | 101 (48.8) |
| ……2. alveolar ground-glass | 27 (17.1) | 4 (8.2) | 31 (15.0) |
| ……3. multifocal | 22 (13.9) | 2 (4.1) | 24 (11.6) |
| ……4. ground-glass opacities | 45 (28.5) | 5 (10.2) | 50 (24.2) |
| ……5. other | 1 (0.6) | 0 (0.0) | 1 (0.5) |
| …Chest X-Ray 2 | 11 (100.0) | 13 (100.0) | 24 (100.0) |
| ……1. normal | 0 (0.0) | 2 (15.4) | 2 (8.3) |
| ……2. alveolar segmento | 0 (0.0) | 0 (0.0) | 0 (0.0) |
| ……3. multifocal | 0 (0.0) | 1 (7.7) | 1 (4.2) |
| ……4. ground-glass opacities | 10 (90.9) | 8 (61.5) | 18 (75.0) |

| | | | |
|---|---|---|---|
| ......5. other | 1 (9.1) | 2 (15.4) | 3 (12.5) |
| ...Chest CT 1 | 241 (100.0) | 301 (100.0) | 542 (100.0) |
| ......1. normal | 35 (14.5) | 56 (18.6) | 91 (16.8) |
| ......2. alveolar segmento | 15 (6.2) | 19 (6.3) | 34 (6.3) |
| ......3. multifocal | 19 (7.9) | 53 (17.6) | 72 (13.3) |
| ......4. ground-glass opacities | 167 (69.3) | 164 (54.5) | 331 (61.1) |
| ......5. other | 5 (2.1) | 9 (3.0) | 14 (2.6) |
| ...Chest CT 2 | 43 (100.0) | 12 (100.0) | 55 (100.0) |
| ......1. normal | 1 (2.3) | 1 (8.3) | 2 (3.6) |
| ......2. alveolar segmento | 0 (0.0) | 0 (0.0) | 0 (0.0) |
| ......3. multifocal | 0 (0.0) | 1 (8.3) | 1 (1.8) |
| ......4. ground-glass opacities | 41 (95.3) | 10 (83.3) | 51 (92.7) |
| ......5. other | 1 (2.3) | 0 (0.0) | 1 (1.8) |
| ...Chest X-Ray 1 | ... | ... | ... |
| ......plus chest CT 1 | 8 (12.7) | 9 (23.7) | 17 (16.8) |
| ......plus chest CT 2 | 0 (0.0) | 0 (0.0) | 0 (0.0) |
| ......plus chest CT 3 | 3 (4.8) | 0 (0.0) | 3 (3.0) |
| ......plus chest CT 4 | 7 (11.1) | 2 (5.3) | 9 (8.9) |
| ......plus chest CT 5 | 0 (0.0) | 0 (0.0) | 0 (0.0) |
| Laboratory results [num. (%)] | ... | ... | ... |
| ...... [median (IQR)] | ... | ... | ... |
| ...Lymphocyte count | 323 (100.0) | 287 (100.0) | 610 (100.0) |
| ...... | 1369 (995-1827) | 1302 (893-1768) | 1336 (936-1796) |
| ......Lymphocyte <1,500 | 184 (57.0) | 171 (59.6) | 355 (58.2) |
| ...White-cell count | 324 (100.0) | 319 (100.0) | 643 (100.0) |
| ...... | 6900 (5100-9625) | 5300 (4200-6450) | 5900 (4600-8000) |
| ...... <4,500 | 48 (14.8) | 95 (29.8) | 143 (22.2) |
| ...... >10,000 | 73 (22.5) | 19 (6.0) | 92 (14.3) |
| ...Platelet | 324 (100.0) | 318 (100.0) | 642 (100.0) |
| ...... | 211800 (164675-278500) | 2e+05 (166250-245750) | 207050 (166000-257750) |
| ...... <150,000 | 58 (17.9) | 46 (14.5) | 104 (16.2) |
| ...ESR | 206 (100.0) | 163 (100.0) | 369 (100.0) |
| ...... | 47 (27-72) | 25 (14-45) | 37 (20-63) |
| ...... >50 | 93 (45.1) | 33 (20.2) | 126 (34.1) |
| ...Creatinine | 323 (100.0) | 329 (100.0) | 652 (100.0) |
| ...... | 1 (1-1) | 1 (1-1) | 1 (1-1) |
| ...... >1.20 | 53 (16.4) | 30 (9.1) | 83 (12.7) |
| ...Lactate dehydrogenase | 314 (100.0) | 249 (100.0) | 563 (100.0) |
| ...... | 434 (321-650) | 236 (171-327) | 340 (228-532) |
| ...... >400 | 182 (58.0) | 41 (16.5) | 223 (39.6) |
| ...... >600 | 100 (31.8) | 5 (2.0) | 105 (18.7) |
| ...C-reactive protein | 177 (100.0) | 265 (100.0) | 442 (100.0) |
| ...... | 50 (13-113) | 15 (5-42) | 23 (7-73) |
| ...... >10 | 136 (76.8) | 165 (62.3) | 301 (68.1) |
| ...... >60 | 82 (46.3) | 50 (18.9) | 132 (29.9) |
| ...... >100 | 59 (33.3) | 23 (8.7) | 82 (18.6) |
| ...Ferritin | 221 (100.0) | 200 (100.0) | 421 (100.0) |
| ...... | 612 (281-1607) | 428 (200-907) | 514 (231-1152) |
| ...... >500 | 125 (56.6) | 92 (46.0) | 217 (51.5) |
| ...... >600 | 112 (50.7) | 80 (40.0) | 192 (45.6) |
| ...Bilirrubin | 297 (100.0) | 306 (100.0) | 603 (100.0) |
| ...... | 1 (0-1) | 0 (0-1) | 0 (0-1) |
| ...Alanine aminotransferase | 324 (100.0) | 307 (100.0) | 631 (100.0) |
| ...... | 33 (20-58) | 24 (15-40) | 28 (17-48) |
| ...... >41 | 124 (38.3) | 72 (23.5) | 196 (31.1) |
| ...Aspartate aminotransferase | 322 (100.0) | 307 (100.0) | 629 (100.0) |
| <- <- | 34 (23-52) | 25 (19-35) | 29 (21-44) |
| ...... >40 | 126 (39.1) | 52 (16.9) | 178 (28.3) |
| ...Procalcitonin | 0 (0.0) | 6 (100.0) | 6 (100.0) |

| | | | |
|---|---|---|---|
| <- <- | – | 0 (0-0) | 0 (0-0) |
| …D-dimer | 9 (100.0) | 207 (100.0) | 216 (100.0) |
| …… | 1 (1-1) | 0 (0-1) | 0 (0-1) |
| …3 normal biomarkers | 3 (0.9) | 11 (3.2) | 14 (2.1) |
| Treatements [num. (%)] | … | … | … |
| …With hidroxychloroquine | 5 (1.5) | 13 (3.8) | 18 (2.7) |
| ……With IMV | 1 (20.0) | 5 (38.5) | 6 (33.3) |
| ……deceased | 1 (20.0) | 1 (7.7) | 2 (11.1) |
| …Without hidroxychloroquine | 321 (98.5) | 329 (96.2) | 650 (97.3) |
| ……With IMV | 31 (9.7) | 12 (3.6) | 43 (6.6) |
| ……deceased | 62 (19.3) | 19 (5.8) | 81 (12.5) |
| …With lopinavir/ritonavir | 8 (2.5) | 11 (3.2) | 19 (2.8) |
| ……With IMV | 2 (25.0) | 5 (45.5) | 7 (36.8) |
| ……deceased | 3 (37.5) | 1 (9.1) | 4 (21.1) |
| …Without lopinavir/ritonavir | 318 (97.5) | 331 (96.8) | 649 (97.2) |
| ……With IMV | 30 (9.4) | 12 (3.6) | 42 (6.5) |
| ……deceased | 60 (18.9) | 19 (5.7) | 79 (12.2) |
| …With plasma | 35 (10.7) | 10 (2.9) | 45 (6.7) |
| ……With IMV | 11 (31.4) | 2 (20.0) | 13 (28.9) |
| ……deceased | 6 (17.1) | 1 (10.0) | 7 (15.6) |
| …Without plasma | 291 (89.3) | 332 (97.1) | 623 (93.3) |
| ……With IMV | 21 (7.2) | 15 (4.5) | 36 (5.8) |
| ……deceased | 57 (19.6) | 19 (5.7) | 76 (12.2) |
| …With azithromycin | 225 (69.0) | 158 (46.2) | 383 (57.3) |
| ……With IMV | 22 (9.8) | 10 (6.3) | 32 (8.4) |
| ……deceased | 40 (17.8) | 7 (4.4) | 47 (12.3) |
| …Without azithromycin | 101 (31.0) | 184 (53.8) | 285 (42.7) |
| ……With IMV | 10 (9.9) | 7 (3.8) | 17 (6.0) |
| ……deceased | 23 (22.8) | 13 (7.1) | 36 (12.6) |
| …With steroids | 114 (35.0) | 86 (25.1) | 200 (29.9) |
| ……With IMV | 24 (21.1) | 12 (14.0) | 36 (18.0) |
| ……deceased | 33 (28.9) | 10 (11.6) | 43 (21.5) |
| …Without steroids | 212 (65.0) | 256 (74.9) | 468 (70.1) |
| ……With IMV | 8 (3.8) | 5 (2.0) | 13 (2.8) |
| ……deceased | 30 (14.2) | 10 (3.9) | 40 (8.5) |
| Evolution [num. (%)] | … | … | … |
| …Temperature >38 | 27 (8.3) | 52 (15.2) | 79 (11.8) |
| …RR >30? | 47 (14.4) | 15 (4.4) | 62 (9.3) |
| <- SAT <93? | 101 (31.0) | 51 (14.9) | 152 (22.8) |
| …IMV | 32 (9.8) | 17 (5.0) | 49 (7.3) |
| …NIV | 0 (0.0) | 3 (0.9) | 3 (0.4) |
| …infiltrados <48 hs | 21 (6.4) | 13 (3.8) | 34 (5.1) |
| …Altered conciousness | 15 (4.6) | 1 (0.3) | 16 (2.4) |
| …Hemodynamic instability | 17 (5.2) | 5 (1.5) | 22 (3.3) |
| …CURB >=2 | 6 (1.8) | 7 (2.0) | 13 (1.9) |
| …ICU admission | 50 (15.3) | 27 (7.9) | 77 (11.5) |
| …IMV prone position | 17 (5.2) | 4 (1.2) | 21 (3.1) |
| …Haemodyalisis | 0 (0.0) | 0 (0.0) | 0 (0.0) |
| …Arrhythmia | 4 (1.2) | 0 (0.0) | 4 (0.6) |
| …Myocarditis/IM | 0 (0.0) | 0 (0.0) | 0 (0.0) |
| …Inotropic agents | 16 (4.9) | 2 (0.6) | 18 (2.7) |
